# Impact of structured medication reviews on prescribing in English Primary Care: a nationwide observational cohort study

**DOI:** 10.1101/2025.07.23.25332062

**Authors:** James P Sheppard, Paul A Bateman, Cynthia Wright-Drakesmith, Christopher Clark, Rebecca K Barnes, Andrew Clegg, Gary A Ford, Seema Gadhia, William Hinton, FD Richard Hobbs, Sundus Jawad, Kamlesh Khunti, Gregory Y. H. Lip, Simon de Lusignan, Jonathan Mant, Deborah McCahon, Bernardo Meza-Torres, Rupert A Payne, Rafael Perera-Salazar, Claire Reidy, Anna Seeley, Samuel Seidu, Katherine Tucker, Rik van der Veen, Marney Williams, Richard J McManus

## Abstract

**Objectives:** The present study aimed to evaluate the impact of structured medication reviews (SMRs), by examining the proportion of eligible patients who received a review in the first two years of the programme, and whether SMRs were associated with changes in prescribing.

**Design:** Retrospective observational cohort study.

**Setting:** Patients registered to primary care practices in England contributing data to the Oxford Clinical Informatics Digital Hub (ORCHID) were included between 1^st^ April 2020 and 30^th^ September 2022.

**Participants:** De-identified data were extracted from the electronic health records of individuals registered to ORCHID practices aged ≥65 years, prescribed one or more medications and fulfilling the specific eligibility criteria for a SMR.

**Main outcome measures:** The primary outcome was the proportion of patients who received a review. Further outcomes included the proportion of potentially inappropriate drug combinations corrected following an SMR. The association between SMRs and prescription changes and primary care contacts was examined by matching individuals who received an SMR to individuals who did not receive an SMR, according to age, sex and primary care practice using cumulative density sampling. Analyses were undertaken using adjusted logistic regression.

**Results:** From a total of 635,698 eligible patients, 82,285 patients (12.94%, 95% confidence interval [CI] 12.86% to 13.02%) received at least one SMR during the study observation period. In those prescribed potentially inappropriate drug combinations prior to an SMR, between 12.5% and 40.0% were corrected up to three months later. In matched analyses, SMRs were associated with a significant increase new prescriptions of ACE inhibitors (adjusted odds ratio [aOR] 1.56, 95%CI 1.35-1.81), statins (aOR 1.78, 95%CI 1.57-2.02), inhaled corticosteroids (aOR 1.19 95%CI 1.05-1.36), opioids (aOR 1.31 95%CI 1.20-1.42), and antidepressants (aOR 1.45 95%CI 1.28-1.63). In those previously prescribed treatment, individuals receiving an SMR were significantly more likely stop ACE inhibitors (aOR 1.37, 95%CI 1.18-1.58), statins (aOR 1.35, 95%CI 1.17-1.56) and antidepressants (antidepressants aOR 1.37 95%CI 1.21-1.56). SMRs were associated with a significant increase in primary care contacts of 0.14 (95% CI 0.13 to 0.16; equivalent to 14 extra patient contacts for every 100 individuals receiving an SMR).

**Conclusions:** SMRs were associated with starting new medications and stopping existing prescriptions compared to usual care. It was unclear if such activity was appropriate or represented improved patient care. Further work is needed to understand if these changes in prescribing improved patient outcomes.

**What is already known on this topic:**

- Inappropriate polypharmacy can expose the most vulnerable patients to decreased quality of life and adverse drug events.
- Outside of trials, studies of pharmacist-led medication reviews undertaken in routine clinical practice have shown little impact on prescribing and patient-centred care.
- Structured medication reviews are a National Institute for Health and Care Excellence (NICE) approved clinical intervention to address complex or problematic polypharmacy and were introduced widely in the UK NHS in 2020.

**What this study adds:**

- We found that one in eight eligible patients received a structured medication review during the first two years of the programme’s rollout in England.
- Structured medication reviews were associated with an increased likelihood of starting medication in those not previously prescribed treatment, and an increased likelihood of stopping medications in those with existing prescriptions.
- This analysis was limited by the data available within primary care electronic health records and so it is unclear if the observed changes in prescribing resulted in improvements in patient outcomes

## Introduction

Prescribing medicines is the most common intervention in the NHS, and most prescribing takes place in primary care.^1,2^ However, inappropriate polypharmacy (where medicines are no longer appropriate)^3^ can expose the most vulnerable patients to decreased quality of life^4–6^ and hospitalisation with adverse drug events.^7–9^ It is estimated that £400 million is spent each year on admissions to hospital caused by harm from potentially inappropriate medication prescriptions.^10^

Over the past twenty years, initiatives have sought to reduce polypharmacy related harm in UK primary care,^11^ and the National Institute for Health and Care Excellence have recommended Structured Medication Reviews (SMR) as a clinical intervention to address complex or problematic polypharmacy.^12^ These were introduced via Primary Care Networks (PCNs) in 2020,^13^ and were designed to be undertaken by clinical pharmacists embedded in primary care, reviewing medications prescribed to patients most at risk of medicines-related harm.^12,13^

Clinical trials focused on chronic diseases have shown that structured medication reviews can improve cardiovascular risk management. However, these studies vary widely in intervention types and measured outcomes.^14^ Furthermore, there is limited evidence that medication reviews reduce adverse drug events.^15,16^ Evaluation studies of pharmacist-led medication reviews undertaken in routine clinical practice have shown mixed results in terms of prescribing and patient-centred care.^17–19^ One possible reason for this is that historically, primary care practices have prioritised being time-efficient rather than being comprehensive, so that most medication reviews were carried out with little or no patient involvement, and medicines were rarely stopped or reduced.^20^ In an early qualitative evaluation undertaken during the first year following the introduction of SMRs, it was suggested that the implementation of the service had been sub-optimal, failing to match the aspiration for patients that was presented in the original policy.^21^

To date, quantitative evaluations of the service have been limited to descriptive analyses of who received an SMR during the COVID-19 pandemic.^22^ The impact of the SMR programme on prescribing in primary care remains unknown. The present study therefore, aimed to evaluate the impact of SMRs on prescribing in primary care by determining the proportion of eligible patients that received an SMR in the first two years and whether SMRs were associated with changes in prescribing and primary care contacts during this period.

## Methods

### Setting

This was a retrospective observational cohort study design, using routine data from primary care electronic health records (EHRs) from GP practices in England collected via the Oxford Clinical Informatics Digital Hub (ORCHID).^23^ Data were extracted from EHRs for the observation period between 1^st^ January 2020 and 31^st^ December 2022. The study received ethical approval from South Central – Hampshire A Research Ethics Committee (ref: 22/SC/0373).

### Population

De-identified data were extracted from the EHRs of patients registered at primary care practices in England contributing data to ORCHID,^23^ who were aged 65 years or over, prescribed one or more repeat medications and who fulfilled at least one of the eligibility criteria for a structured medication review, as defined in the PCN contract for SMRs.^24^ Specifically, these were individuals who were either in a care home, had complex polypharmacy (prescribed 10 or more medications), were prescribed medications commonly associated with medication errors (see table S1), had severe frailty (defined as an electronic Frailty Index^25^ of >0.36) or were prescribed one or more potentially addictive medications (i.e. opioids, gabapentin, benzodiazepines or non-benzodiazepine hypnotic medications [z-drugs]).^24^ Individuals entered the cohort on the 1^st^ January 2020 and SMRs that occurred between 1^st^ April 2020 and 30^th^ September 2022 were included. Data from the three months prior to the SMR observation period (January-March 2020) were used to define treatment prescriptions. Medication changes (new prescriptions and stoppages of existing prescriptions) up to three months after an SMR were included up until 31^st^ December 2022.

### Outcomes

The primary outcome of this study was the proportion of eligible patients receiving an SMR during the SMR observation period. Secondary outcomes included changes in medication prescriptions (new prescriptions and stoppages of existing prescriptions) following an SMR. Analyses focussed on prescription of blood pressure lowering drugs (ACE inhibitors, alpha blockers, angiotensin II receptor blockers, beta blockers, calcium channel blockers, thiazide and thiazide-like diuretics), other cardiovascular prevention medications (statins, antiplatelets, direct oral anticoagulants [DOACs], vitamin K antagonists [VKA]), inhaled medications (Inhaled beta agonist, Inhaled corticosteroids, not including antimuscarinic drugs [tiotropium/ipratropium used in asthma/COPD]), pain medications (NSAIDS, opioids), psychotropic medications (antidepressants, benzodiazepines, gabapentin, pregabalin, Z-drugs) and other treatments of interest including donepezil, laxatives and proton pump inhibitors (PPIs). Further analyses focussed on changes in medications identified in the PCN contract as being potentially inappropriate (see table S1 for details). Finally, changes in any primary care contacts between healthcare professionals and patients before and after an SMR were examined. For this analysis, healthcare professionals were defined as general practitioners (GP), pharmacists, Health Care Assistants (HCA) or nurses. Administrative staff were not included. Face-to-face and remote (telephone, video calls, etc) patient contacts were not differentiated as these data were often missing and not consistently defined.

### Exposures

The primary outcome (prevalence of SMRs) was descriptive and so no exposure was defined. For secondary outcomes (changes in medication prescription and primary care contacts), the exposure of interest was an SMR during the observation period 1^st^ April 2020 to 30^th^ September 2022. SMRs were defined using the Systematized Nomenclature of Medicine (SNOMED) clinical term 1239511000000100 which was defined in the PCN contract.

### Covariates

All analyses examining the association between SMRs and outcomes were adjusted for covariates defined according to data available at any timepoint prior to the observation period (i.e., before April 2020) and included body mass index (BMI) ethnicity, indices of multiple deprivation, smoking status, care home residence, baseline cholesterol, electronic frailty index score^25^ and number of multiple long-term conditions. Age (defined at the time of the index date) and sex were not included in the modelling as these were used as matching variables for the cohort. The latter were defined based on the list of 37 conditions included in the Cambridge Multimorbidity Score.^26,27^

### Statistical analysis

Descriptive analyses were used to determine the proportion of patients potentially eligible to receive an SMR during the study observational period. The characteristics of this population were categorised by whether or not an individual received an SMR. In those who received an SMR and had continuous follow-up (i.e., those who survived and remained within ORCHID-registered practices so that medication prescription could be measured) during the observation period, changes in medication prescription (stopped or started) within 3 months (91 days) of an SMR were examined according to individual drug class. Furthermore, potentially inappropriate medication prescriptions were examined by identifying individuals with specific conditions or concurrent drug prescriptions which would put them at risk of adverse drug events, as detailed in the PCN contract for SMRs (see table S1 for details).^28^ The proportion of these potential medication errors that were corrected within 3 months of an SMR was estimated using descriptive statistics.

To examine the associations between SMRs and prescription changes and primary care contacts, a cohort of patients who received an SMR were matched to controls who did not receive an SMR using cumulative density sampling. Individuals were matched 1:1 according to baseline age (5-year groups), sex and primary care practice. Each control was given an index date which was the date their matched case received an SMR. Analyses of the association between SMRs and outcomes were undertaken using logistic regression models, which were adjusted baseline covariates (listed above), with missing data for index of multiple deprivation dealt with using multiple imputation (10 imputations). For other binary or categorical variables included in the modelling, the proportion of missing data were less than 5% so analyses were restricted to individuals with complete data. All analyses were undertaken using Stata version 18.0 (StataCorp, College Station, Texas, USA).

### Patient and public involvement

This study was developed and conducted with the help of our patient and public advisor Marney Williams. As a member of our study advisory group, they commented on the study protocol and have been present in all team meetings discussing results and reporting.

## Results

A total of 635,698 patients, from 783 practices across England were identified as being eligible for an SMR. Of these, 82,285 patients (12.94%, 95% confidence interval [CI] 12.86% to 13.02%) received at least one SMR during the study observational period. More patients received an SMR in the second year of the observation period than the first year (figure S1). Patients who received an SMR were on average aged 77 years, 59.6% female, with 3.3% Asian ethnicity and 1.6% black ethnicity. Compared to those who did not receive an SMR, there were higher proportions of residents in nursing homes, moderate to severe frailty, hyper-polypharmacy (≥10 prescriptions), and complex multiple long-term conditions (≥4 conditions present) in patients who did receive one (tables 1, S2 and S3). There were no observed differences in the proportion of patients receiving an SMR on the basis of age, sex, ethnicity or social deprivation (table 1).

**Table 1.**
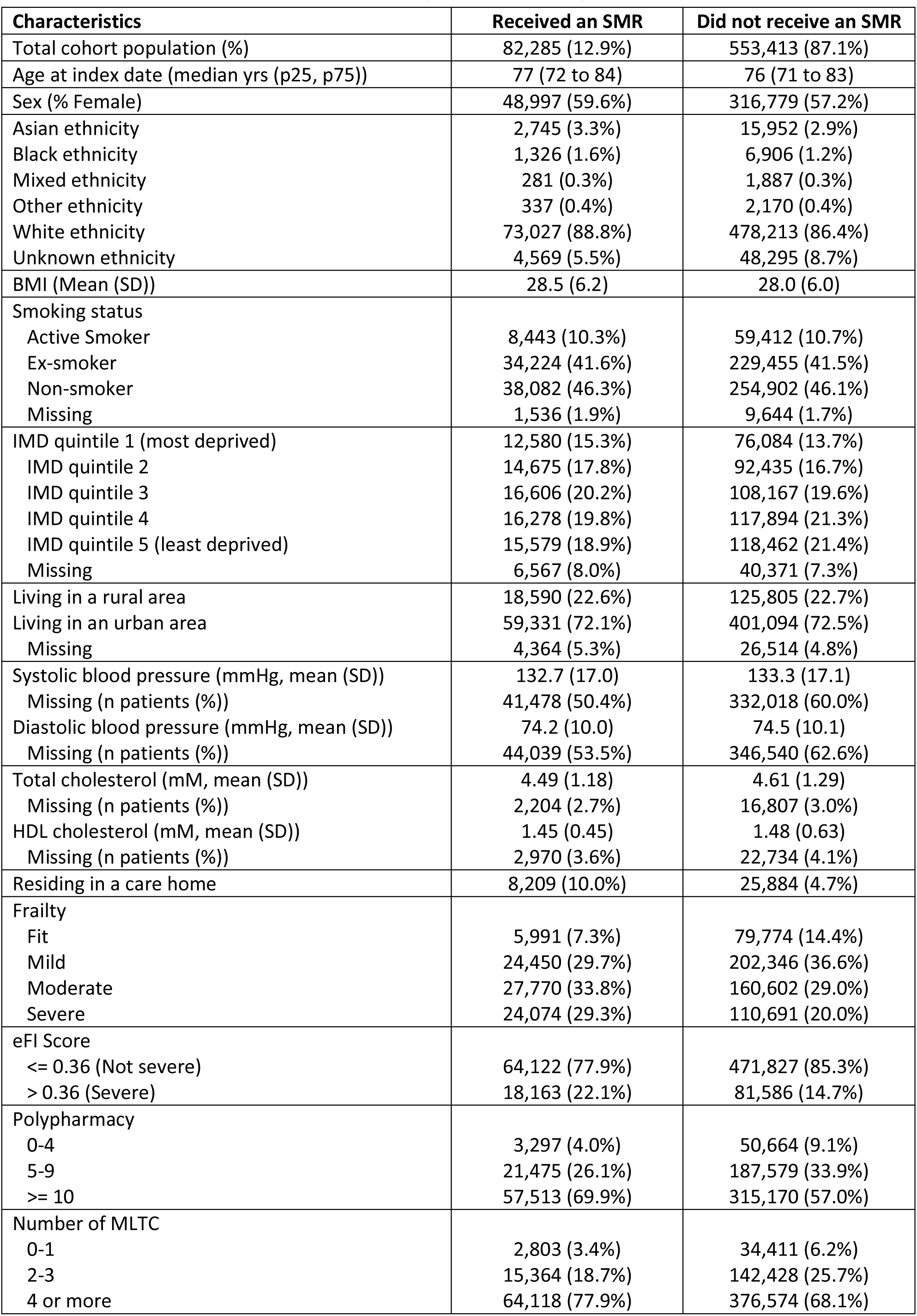
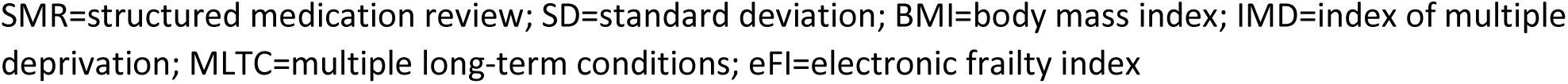
Baseline characteristics of those receiving and not receiving a structured medication review.

| Characteristics | Received an SMR | Did not receive an SMR |
| --- | --- | --- |
| Total cohort population (%) | 82,285 (12.9%) | 553,413 (87.1%) |
| Age at index date (median yrs (p25, p75)) | 77 (72 to 84) | 76 (71 to 83) |
| Sex (% Female) | 48,997 (59.6%) | 316,779 (57.2%) |
| Asian ethnicity | 2,745 (3.3%) | 15,952 (2.9%) |
| Black ethnicity | 1,326 (1.6%) | 6,906 (1.2%) |
| Mixed ethnicity | 281 (0.3%) | 1,887 (0.3%) |
| Other ethnicity | 337 (0.4%) | 2,170 (0.4%) |
| White ethnicity | 73,027 (88.8%) | 478,213 (86.4%) |
| Unknown ethnicity | 4,569 (5.5%) | 48,295 (8.7%) |
| BMI (Mean (SD)) | 28.5 (6.2) | 28.0 (6.0) |
| Smoking status |  |  |
| Active Smoker | 8,443 (10.3%) | 59,412 (10.7%) |
| Ex-smoker | 34,224 (41.6%) | 229,455 (41.5%) |
| Non-smoker | 38,082 (46.3%) | 254,902 (46.1%) |
| Missing | 1,536 (1.9%) | 9,644 (1.7%) |
| IMD quintile 1 (most deprived) | 12,580 (15.3%) | 76,084 (13.7%) |
| IMD quintile 2 | 14,675 (17.8%) | 92,435 (16.7%) |
| IMD quintile 3 | 16,606 (20.2%) | 108,167 (19.6%) |
| IMD quintile 4 | 16,278 (19.8%) | 117,894 (21.3%) |
| IMD quintile 5 (least deprived) | 15,579 (18.9%) | 118,462 (21.4%) |
| Missing | 6,567 (8.0%) | 40,371 (7.3%) |
| Living in a rural area | 18,590 (22.6%) | 125,805 (22.7%) |
| Living in an urban area | 59,331 (72.1%) | 401,094 (72.5%) |
| Missing | 4,364 (5.3%) | 26,514 (4.8%) |
| Systolic blood pressure (mmHg, mean (SD)) | 132.7 (17.0) | 133.3 (17.1) |
| Missing (n patients (%)) | 41,478 (50.4%) | 332,018 (60.0%) |
| Diastolic blood pressure (mmHg, mean (SD)) | 74.2 (10.0) | 74.5 (10.1) |
| Missing (n patients (%)) | 44,039 (53.5%) | 346,540 (62.6%) |
| Total cholesterol (mM, mean (SD)) | 4.49 (1.18) | 4.61 (1.29) |
| Missing (n patients (%)) | 2,204 (2.7%) | 16,807 (3.0%) |
| HDL cholesterol (mM, mean (SD)) | 1.45 (0.45) | 1.48 (0.63) |
| Missing (n patients (%)) | 2,970 (3.6%) | 22,734 (4.1%) |
| Residing in a care home | 8,209 (10.0%) | 25,884 (4.7%) |
| Frailty |  |  |
| Fit | 5,991 (7.3%) | 79,774 (14.4%) |
| Mild | 24,450 (29.7%) | 202,346 (36.6%) |
| Moderate | 27,770 (33.8%) | 160,602 (29.0%) |
| Severe | 24,074 (29.3%) | 110,691 (20.0%) |
| eFI Score |  |  |
| <= 0.36 (Not severe) | 64,122 (77.9%) | 471,827 (85.3%) |
| > 0.36 (Severe) | 18,163 (22.1%) | 81,586 (14.7%) |
| Polypharmacy |  |  |
| 0-4 | 3,297 (4.0%) | 50,664 (9.1%) |
| 5-9 | 21,475 (26.1%) | 187,579 (33.9%) |
| >= 10 | 57,513 (69.9%) | 315,170 (57.0%) |
| Number of MLTC |  |  |
| 0-1 | 2,803 (3.4%) | 34,411 (6.2%) |
| 2-3 | 15,364 (18.7%) | 142,428 (25.7%) |
| 4 or more | 64,118 (77.9%) | 376,574 (68.1%) |

SMR=structured medication review; SD=standard deviation; BMI=body mass index; IMD=index of multiple deprivation; MLTC=multiple long-term conditions; eFI=electronic frailty index

The majority of patients had no changes in their treatment following a SMR (table 2). In those already prescribed treatment, the medications most commonly stopped were NSAIDs (29.6% stopped), benzodiazepines (22.3% stopped), laxatives (19.9% stopped) and Z-drugs (16.1% stopped) and opioids (15.5% stopped). In patients not previously taking a given drug therapy, PPIs (11.3% started), statins (7.7% started), opioids (7.6% started) and laxatives (6.4%) were most likely to be started, although the proportion starting treatment was smaller (compared to the proportion stopping) (table 2). With regard to potentially inappropriate prescriptions, between 12.5% and 40.0% were corrected, although the overall numbers of inappropriate prescriptions were small (table 3). Medication combinations that increase the risk of a gastrointestinal bleeds were most commonly corrected (between 31.2 and 40.0% of combinations corrected), whereas drugs that could exacerbate asthma (12.5% corrected) or lead to respiratory depression (14.5% corrected) were least commonly corrected.

**Table 2.**
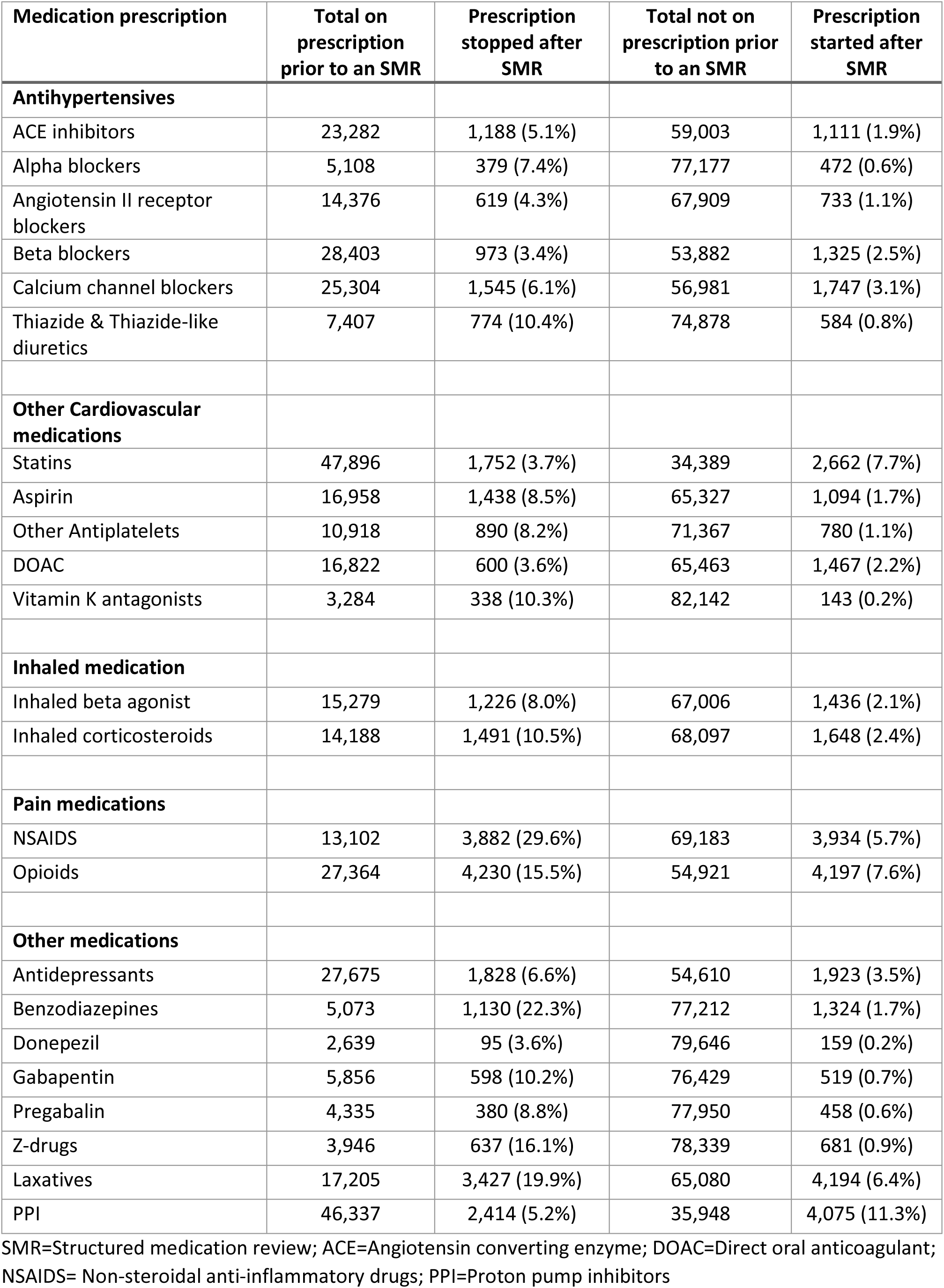
Changes in medication prescription following a structure medication review (n=82,285)

| Medication prescription | Total on prescription prior to an SMR | Prescription stopped after SMR | Total not on prescription prior to an SMR | Prescription started after SMR |
| --- | --- | --- | --- | --- |
| **Antihypertensives** |  |  |  |  |
| ACE inhibitors | 23,282 | 1,188 (5.1%) | 59,003 | 1,111 (1.9%) |
| Alpha blockers | 5,108 | 379 (7.4%) | 77,177 | 472 (0.6%) |
| Angiotensin II receptor blockers | 14,376 | 619 (4.3%) | 67,909 | 733 (1.1%) |
| Beta blockers | 28,403 | 973 (3.4%) | 53,882 | 1,325 (2.5%) |
| Calcium channel blockers | 25,304 | 1,545 (6.1%) | 56,981 | 1,747 (3.1%) |
| Thiazide & Thiazide-like diuretics | 7,407 | 774 (10.4%) | 74,878 | 584 (0.8%) |
| **Other Cardiovascular medications** |  |  |  |  |
| Statins | 47,896 | 1,752 (3.7%) | 34,389 | 2,662 (7.7%) |
| Aspirin | 16,958 | 1,438 (8.5%) | 65,327 | 1,094 (1.7%) |
| Other Antiplatelets | 10,918 | 890 (8.2%) | 71,367 | 780 (1.1%) |
| DOAC | 16,822 | 600 (3.6%) | 65,463 | 1,467 (2.2%) |
| Vitamin K antagonists | 3,284 | 338 (10.3%) | 82,142 | 143 (0.2%) |
| **Inhaled medication** |  |  |  |  |
| Inhaled beta agonist | 15,279 | 1,226 (8.0%) | 67,006 | 1,436 (2.1%) |
| Inhaled corticosteroids | 14,188 | 1,491 (10.5%) | 68,097 | 1,648 (2.4%) |
| **Pain medications** |  |  |  |  |
| NSAIDS | 13,102 | 3,882 (29.6%) | 69,183 | 3,934 (5.7%) |
| Opioids | 27,364 | 4,230 (15.5%) | 54,921 | 4,197 (7.6%) |
| **Other medications** |  |  |  |  |
| Antidepressants | 27,675 | 1,828 (6.6%) | 54,610 | 1,923 (3.5%) |
| Benzodiazepines | 5,073 | 1,130 (22.3%) | 77,212 | 1,324 (1.7%) |
| Donepezil | 2,639 | 95 (3.6%) | 79,646 | 159 (0.2%) |
| Gabapentin | 5,856 | 598 (10.2%) | 76,429 | 519 (0.7%) |
| Pregabalin | 4,335 | 380 (8.8%) | 77,950 | 458 (0.6%) |
| Z-drugs | 3,946 | 637 (16.1%) | 78,339 | 681 (0.9%) |
| Laxatives | 17,205 | 3,427 (19.9%) | 65,080 | 4,194 (6.4%) |
| PPI | 46,337 | 2,414 (5.2%) | 35,948 | 4,075 (11.3%) |

SMR=Structured medication review; ACE=Angiotensin converting enzyme; DOAC=Direct oral anticoagulant; NSAIDS= Non-steroidal anti-inflammatory drugs; PPI=Proton pump inhibitors

**Table 3.** Changes in potentially inappropriate medication prescriptions following a structured medication review.

| Indicator | Condition | Drug or drug combination | Total with inappropriate medication combinations | Not corrected following SMR (n(%)) | Corrected following SMR (n(%)) |
| --- | --- | --- | --- | --- | --- |
| GIB01 | GI bleed | NSAID, but not PPI | 101 | 65 (64.4%) | 36 (35.6%) |
| GIB02 | GI bleed | NSAID & anticoagulant | 190 | 114 (60.0%) | 76 (40.0%) |
| GIB03 | GI bleed | NSAID & anticoagulant, but not PPI | 35 | 27 (77.1%) | 8 (40.0%) |
| GIB04 | GI bleed | Aspirin & Antiplatelet, but not PPI | 93 | 64 (68.8%) | 29 (31.2%) |
| GIBCI | GI bleed | Unique patients from GIB01 to GIB04 | 324 | 201 (62.0%) | 123 (34.0%) |
| PAIN01 | Respiratory depression/confusion | Opioid & Benzo/Gabapentin/Pregabalin/Z-drug | 72 | 57 (79.2%) | 15 (20.8%) |
| PAIN02 | Constipation | Opioid, but not laxative | 1,001 | 766 (76.5%) | 235 (23.5%) |
| PAIN03 | Respiratory depression/confusion | Opioid | 202 | 172 (85.1%) | 30 (14.9%) |
| FRAC01a | Fall | Z-drug | 1,218 | 1002 (82.3%) | 216 (17.7%) |
| FRAC01b | Fracture | Z-drug | 276 | 226 (81.9%) | 50 (18.1%) |
| FRAC02a | Fall | Benzo | 1,741 | 1295 (74.4%) | 446 (25.6%) |
| FRAC02b | Fracture | Benzo | 456 | 331 (72.6%) | 125 (27.4%) |
| FRAC03a | Fall | Benzo/Z-drug | 191 | 119 (62.3%) | 72 (37.7%) |
| FRAC03b | Fracture | Benzo/Z-drug | 687 | 480 (69.9%) | 207 (30.1%) |
| RESP01 | Asthma | LABA, but not inhaled corticosteroid | 112 | 98 (87.5%) | 14 (12.5%) |

GI=gastro-intestinal bleed; NSAID=non-steroidal anti-inflammatory drug; PPI=proton pump inhibitor; SMR=Structured medication review; DOAC=direct oral anticoagulant; LABA=inhaled Long Acting Beta-agonist

In 71,939 patients who received an SMR, matched to 71,939 patients who did not (table S4), medication reviews were associated with a significant increase in starting new medications and stopping existing prescriptions across nearly all drug classes of interest. This included increased prescribing of antihypertensives, cardiovascular, inhaled, pain, psychotropic, and other medications in patients receiving an SMR (figure 1). Furthermore, those already prescribed antihypertensives, cardiovascular, psychotropic, laxatives and PPIs were more likely to have them stopped if they received an SMR (figure 2). There was no evidence of an association between SMRs and stopping inhaled or pain medications, with the exception of NSAIDs (figure 2).

**Figure 1.**
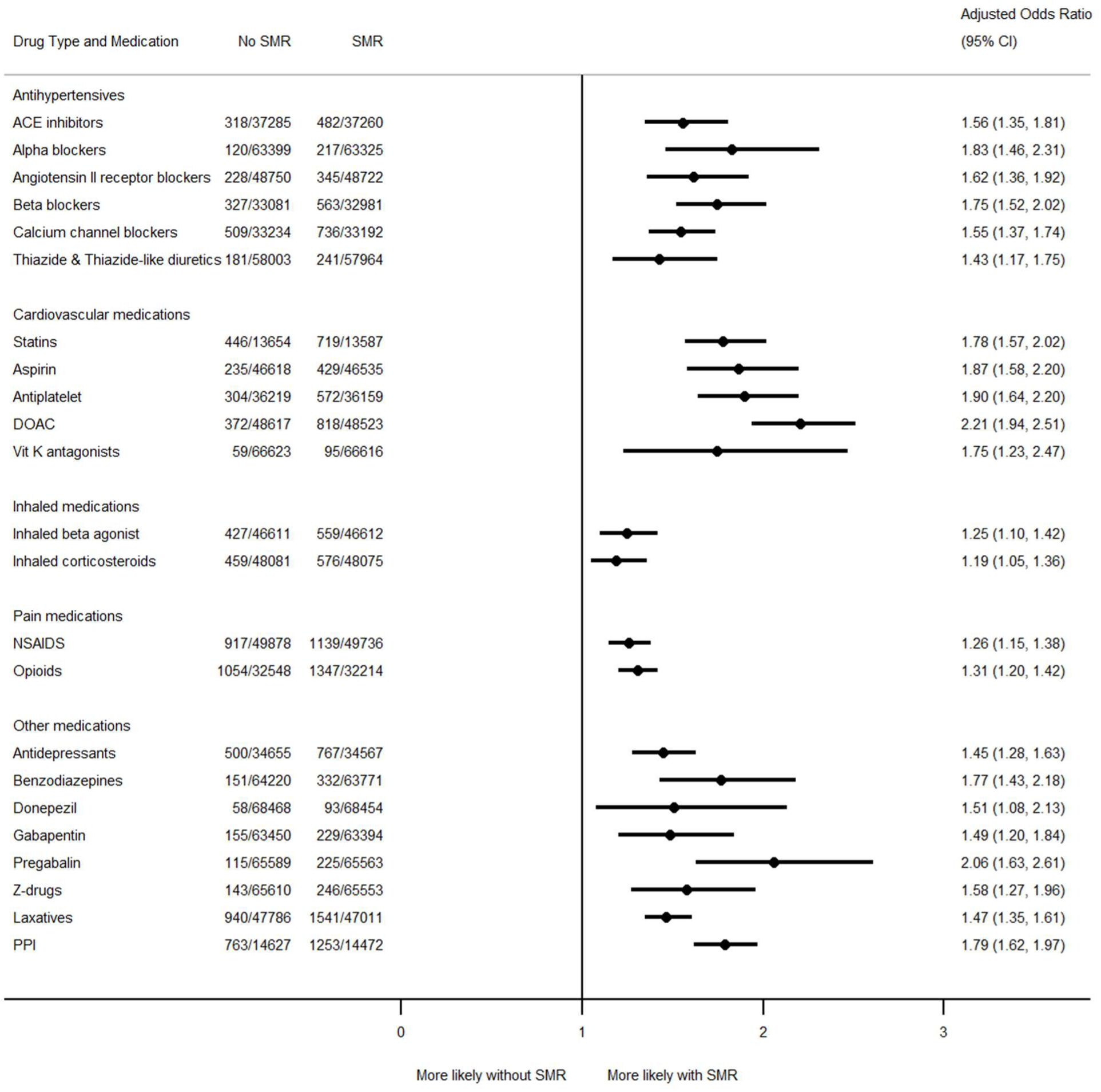
Association between structured medication reviews and starting medications for the first time. Models adjusted for body mass index, ethnicity, indices of multiple deprivation, smoking status, care home residence, baseline cholesterol, electronic frailty index score^25^ and number of multiple long-term conditions.^26^ SMR=Structured medication review; CI=Confidence interval; ACE=Angiotensin-converting enzyme; DOAC=Direct oral anticoagulant; Vit K=Vitamin K; NSAIDS= Non-steroidal anti-inflammatory drugs; PPI=Proton pump inhibitors

**Figure 2.**
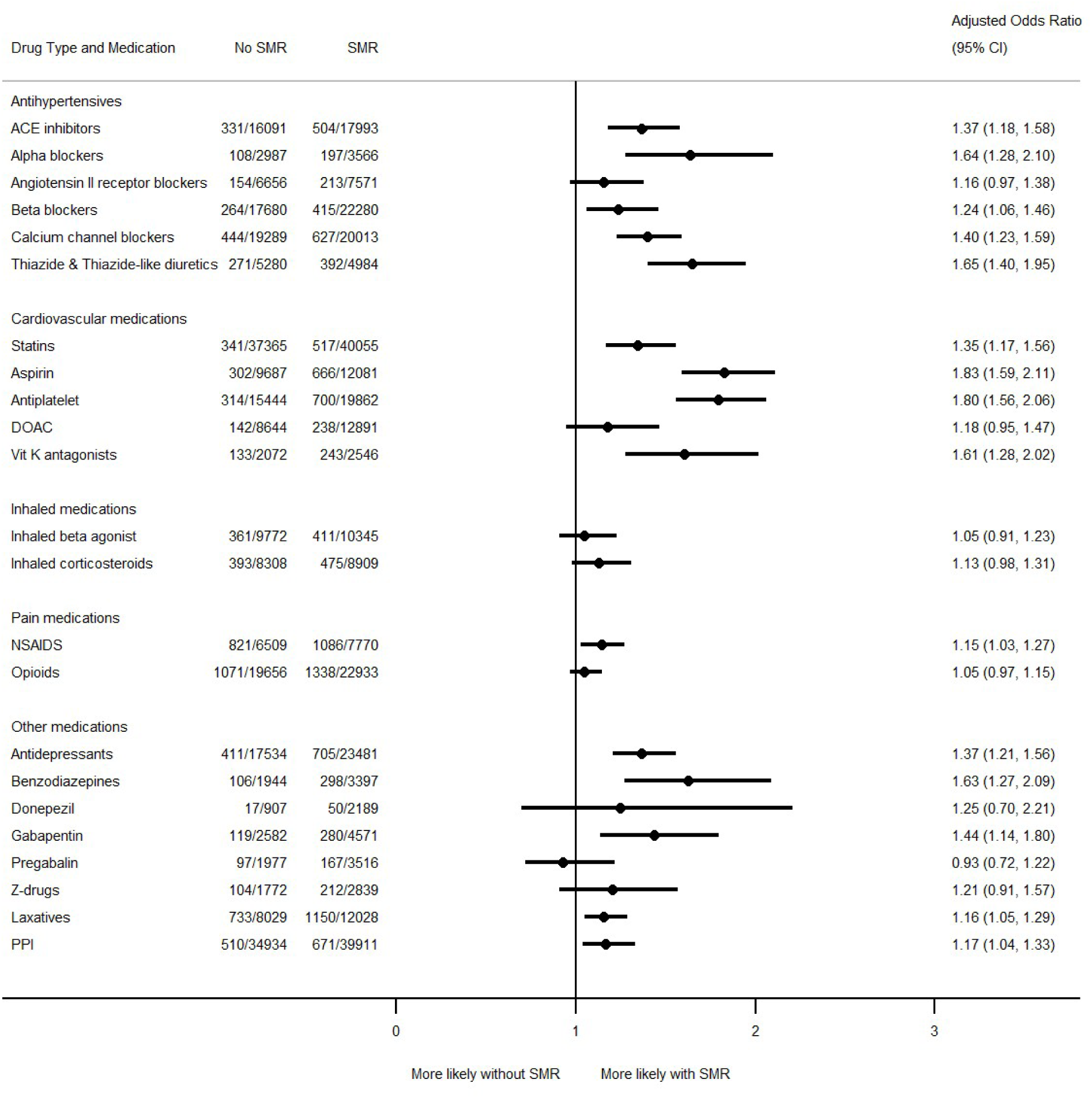
Association between structured medication reviews and stopping medications prescribed prior to the index date. Models adjusted for body mass index, ethnicity, indices of multiple deprivation, smoking status, care home residence, baseline cholesterol, electronic frailty index score^25^ and number of multiple long-term conditions.^26^ SMR=Structured medication review; CI=Confidence interval; ACE=Angiotensin converting enzyme; DOAC=Direct oral anticoagulant; Vit K=Vitamin K; NSAIDS= Non-steroidal anti-inflammatory drugs; PPI=Proton pump inhibitors

Patients receiving a structured medication review had an average of 4 (IQR 2-7) primary care contacts with clinical personnel in the three months prior to their review, and 5 (IQR 3-8) contacts in the three months after. In contrast, those not receiving an SMR had an average of 3 (IQR 1-5) contacts before and after the index date, meaning that SMRs were associated with a significant increase in primary care contacts of 0.14 (95% CI 0.13 to 0.16; equivalent to 14 extra patient contacts for every 100 individuals receiving an SMR).

Results were similar in analyses using different definitions to describe primary care contacts, including those where contacts with a code for an SMR were excluded (table S5).

## Discussion

### Summary of main findings

In this analysis, one in eight eligible patients received an SMR in the first two years of the SMR service rollout in England. This reflected a time of significant change for the UK National Health Service as initial implementation coincided with the first winter of the COVID-19 pandemic alongside major changes in staffing including a large increase in the numbers of pharmacists employed in primary care.^29^ SMRs that were undertaken appeared appropriately targeted toward patients with greater frailty and multiple long-term conditions, without any clear demographic or socioeconomic bias.

As might be expected, many people continued with their prescribed medication following an SMR, but modest net changes in prescribing masked a significant increase in starting new medications and stopping existing prescriptions across nearly all drug classes of interest. In particular, SMRs were associated with an increased likelihood of stopping most antihypertensive medication classes, statins, antiplatelets, NSAIDs, and some psychotropic medications such as antidepressants, benzodiazepines and gabapentin. However, other psychotropic medications, such as pregabalin and z-drugs, and pain medications such as opioids were less likely to be stopped following an SMR, reflecting clinical caution or patient resistance to discontinuing medications that are prescribed for persistent symptoms despite limited evidence for long-term benefit.

Although this analysis showed that SMRs are associated with increases in starting new medications and stopping of existing prescriptions, it remains unclear if these observed changes were suitable or resulted in a reduction in inappropriate polypharmacy. Structured medication reviews are complex interventions, and so further work is needed to understand the patient and healthcare professional experience of SMRs, as well as whether the changes in prescribing improved patient outcomes in the longer-term.

### Strengths and limitations

This was a large nationwide observational study including SMRs undertaken across England covering the first two years of the service. The large sample size available meant that medication prescription changes before and after an SMR could be examined across a range of drug classes. However, the present analyses were unable to take into account changes in dosing, likely to be an important aspect of an SMR, which tend to be recorded in free text fields which were not available for this study. Furthermore, other holistic care activities over and above prescribing were not captured in this analysis, such as addressing patient or carer medicines-related concerns or issues with medicines self-management, supporting medicines literacy, giving lifestyle advice, sign posting to other services and supporting continuity and coordination of care to sustain safe and appropriate medicines use.

Analyses of inappropriate medication indicators included prescriptions of certain drug combinations, in the presence of medical conditions that would make prescription inappropriate. Many of these scenarios required information about whether an individual had recently attended hospital with an acute event (e.g. a fall or gastrointestinal bleed). However, since linked hospital data were not available, and information about acute events occurring in secondary care is not well coded in GP records,^30,31^ it was not possible to accurately determine whether certain medication combinations were inappropriate or not, and this aspect of the analysis should be interpreted with caution. Furthermore, these analyses focussed only on individuals who received an SMR and it was not possible to examine whether the observed changes in prescribing were sustained over time or resulted in improved patient outcomes (e.g., a reduction in falls, fractures, gastrointestinal bleeds, all-cause hospitalisation or mortality).

The before and after study design defined medications changes in the three months before and three months after an SMR. This was chosen as representing the longest time period that repeat prescriptions of medications for long term conditions are likely to be prescribed.^32^ However, medication prescribing patterns are rarely uniform and therefore defining the exposures using shorter or longer time frames may have resulted in slightly different findings.

### Comparison with previous literature

Few studies have sought to determine the impact of structured medication reviews on prescribing practices in primary care. The present study observed that 13% of eligible patients received an SMR within the first two years of the programme. One previous study using data from routine electronic health record data from the OpenSafely database observed a lower percentage of patients (3.6%) receiving SMRs,^22^ however, that study had a broader focus on all types of medication review, and the denominator population was not limited to patients eligible for an SMR as it was in the present analysis.

Approximately one third of inappropriate medication combinations associated with gastrointestinal bleeding (NSAIDs, anticoagulants, aspirin and antiplatelets but not PPIs) were corrected following an SMR in the present study. Similar findings were observed in a recent study examining the scale-up and rollout of the PINCER intervention, a pharmacist-led approach to reduce hazardous prescribing in primary care.^33^ Across 343 general practices implementing the intervention in routine clinical care, decreases in inappropriate prescribing were observed for all indicators, including a 24% reduction in medication combinations associated with gastrointestinal bleeding.^34^

Evidence from randomised controlled trials suggests that medication reviews result in changes in prescribing and some surrogate clinical endpoints such as blood pressure and cholesterol,^14^ however, there is no evidence that they reduce mortality, hospital admissions, healthcare use, number of patients falling, or improve physical and cognitive functioning and quality of life.^15,16,35^ The present analysis shows that these effects on prescribing could be realised in routine clinical practice as well, however, the effect on clinical and cost-effectiveness outcomes remains unknown and requires further research.

### Implications for clinical practice and policy makers

The present study found evidence that structured medication reviews are associated with significant increases in starting new medications and stopping existing prescriptions, in vulnerable patients with frailty, multiple long-term conditions and polypharmacy. There was evidence that some inappropriate medication combinations, such as those related to an increased risk of gastrointestinal bleeding were corrected following an SMR, although the majority were still prescribed in the three months after an SMR. Addressing such inappropriate prescriptions could have significant consequences for the healthcare system, where recent analyses suggest that inappropriate prescribing of NSAIDs alone (without proton-pump inhibitors) could cost the NHS £2.46 billion per year, with a loss of nearly 2,000 quality adjusted life years.^36^ This needs to be set against the not inconsiderable cost of SMRs: a band 8a practice pharmacist costs £77 per working hour and given preparation and consequential time around an SMR plus the review itself then an hour of time is probably a reasonable estimate.^37^ This means that in the 783 practices assessed 82,285 patients having at least one SMR would have cost at least £6.3m or over £8,000 per practice for 105 SMRs on average.

SMRs were associated with a small increase in primary care contacts, defined as face-to-face or telephone/ video appointments with GPs, pharmacists, HCAs or nurses. Whether this is to be expected is unclear, however, the PCN contract specification for SMRs^28^ does outline expectations for follow-up of SMRs, which could explain the observed increase in primary care contacts.

Despite first being rolled out during the COVID-19 pandemic, 1 in 8 eligible patients received an SMR, with evidence of targeting patients with greater frailty, more polypharmacy and larger numbers of long-term conditions, with no evidence of demographic or socioeconomic bias. This suggests that SMRs were targeted as intended, at those most likely to be at risk of inappropriate polypharmacy and adverse drug events.

However, this study did not examine the impact of SMR related changes in prescribing on hospitalisations or mortality, and so it remains unclear if SMRs improved patient outcomes in the longer-term.

## Conclusions

This study found that 1 in 8 eligible patients received a structured medication review within the first two years of the service rollout. SMRs were associated with a significant increase in starting new medications and stopping existing prescriptions across nearly all drug classes of interest. However, further work is needed to understand whether these changes in prescribing improved patient outcomes in the longer- term.

## Data Availability

Requests for data sharing should be made directly to the Primary Care Hosted Research Datasets Independent Scientific Committee (PrimDISC), based at the University of Oxford. Code lists used to define variables included in the dataset are available at https://github.com/ndpchs-cprd/PD-0002-2022-OSCAR.

## Acknowledgements

We thank Lucy Curtin for administrative support throughout the project. We are very grateful to all those patients who permit their anonymised routine NHS data to be used for this approved research.

## Author contributions

RMc conceived the study and wrote the protocol with KT, JS, RB, RP-S, PB, CW-D, RH, SdeL and GF. CW-D carried out the data management and had full access to the database to create the study population. PB undertook the main analyses. JPS wrote the first draft of the manuscript. All authors revised the manuscript and approved the final version. RMc is the guarantor for this work and accepts full responsibility for the conduct of the study, had access to the data, and controlled the decision to publish. The corresponding author (JPS) attests that all listed authors meet authorship criteria and that no others meeting the criteria have been omitted. JPS and RMc had final responsibility for the decision to submit for publication.

## Funding source

This work was funded by the National Institute for Health Research (NIHR) Applied Research Collaboration Multiple Long-term Conditions Cross-ARC collaboration. JPS received funding from the Wellcome Trust/Royal Society via a Sir Henry Dale Fellowship (ref: 211182/Z/18/Z), and now receives funding via an NIHR Advanced Fellowship (NIHR303621). RB receives funding via an NIHR Advanced Fellowship (NIHR302557). FDRH, RM and KT were supported by NIHR Applied Research Collaboration Oxford and the Thames Valley (OxTV-ARC). RM is an NIHR Senior Investigator.

KK and SS are supported by the NIHR Applied Research Collaboration East Midlands (ARC EM), NIHR Global Research Centre for Multiple Long Term Conditions, NIHR Cross NIHR Collaboration for Multiple Long Term Conditions, NIHR Leicester Biomedical Research Centre (BRC) and the British Heart Foundation (BHF) Centre of Excellence.

The views expressed are those of the author(s) and not necessarily those of the NHS, the NIHR or the Department of Health and Social Care. For the purpose of Open Access, the author has applied a CC BY public copyright licence to any Author Accepted Manuscript version arising from this submission.

## Role of the funding source

The sponsor and funders had no role in the design and conduct of the study; collection, management, analysis, and interpretation of the data; preparation, review, or approval of the manuscript; and decision to submit the manuscript for publication.

## Competing interests declaration

All authors have completed the ICMJE uniform disclosure form at www.icmje.org/coi_disclosure.pdfwww.icmje.org/coi_disclosure.pdf and declare no conflicts of interest.

## Ethics approval

The study protocol was approved by South Central – Hampshire A Research Ethics Committee (ref 22/SC/0373).

## Transparency statement

RM affirms that the manuscript is an honest, accurate, and transparent account of the study being reported; that no important aspects of the study have been omitted; and that any discrepancies from the study as originally planned (and, if relevant, registered) have been explained.

## Data sharing

The Corresponding Author has the right to grant on behalf of all authors and does grant on behalf of all authors, an exclusive licence (or non exclusive for government employees) on a worldwide basis to the BMJ Publishing Group Ltd to permit this article (if accepted) to be published in BMJ editions and any other BMJPGL products and sublicences such use and exploit all subsidiary rights, as set out in our licence.

## Notes

### Competing Interest Statement

The authors have declared no competing interest.

### Author Declarations

The study protocol was approved by South Central Hampshire A Research Ethics Committee (ref 22/SC/0373).

