## Appendix for "Impact of structured medication reviews on prescribing in English Primary Care: a nationwide observational cohort study"

**Supplementary appendix**

James P Sheppard, Paul A Bateman, Cynthia Wright-Drakesmith, Christopher Clark, Rebecca Barnes, Andrew Clegg, Gary Ford, Seema Gadhia, William Hinton, FD Richard Hobbs, Sundus Jawad, Kamlesh Khunti, Gregory Lip, Simon de Lusignan, Jonathan Mant, Deborah McCahon, Bernardo Meza-Torres, Rupert Payne, Rafael Perera-Salazar, Claire Reidy, Anna Seeley, Samuel Seidu, Katherine Tucker, Rik van der Veen, Marney Williams, Richard J McManus

**Contents**

1. **Table S1.** Medication prescriptions commonly associated with medication errors
2. **Figure S1.** Incidence of first structured medication reviews by month of observation
3. **Table S2.** Multiple long-term conditions recorded at baseline in those patients receiving and not receiving a structured medication review
4. **Table S3.** Medications of interest recorded at baseline in those patients receiving and not receiving a structured medication review
5. **Table S4.** Matched cohort baseline characteristics of those receiving and not receiving a structured medication review
6. **Table S5.** Patient contacts three months before and after the index data in those patients receiving and not receiving a structured medication review

**Table S1.** Medication prescriptions commonly associated with medication errors.

| Indicator | Descriptor in Primary Care Network contract | Indicator population in study dataset | Implementation in study dataset |
|---|---|---|---|
| Indicator 1 – GIB01 | Patients 65 years old or over admitted to hospital with a gastro-intestinal (GI) bleed prescribed a non-steroidal anti-inflammatory drug (NSAID) and NOT concurrently prescribed a gastro-protective medicine | Patients 65 years old or over with a primary care code for GI bleed in 5 years prior to first SMR or start of study, prescribed a NSAID and not concurrently prescribed a gastro-protective medicine in the 3 months prior to SMR | In the 3 months following a SMR, prescriptions were classed as changed appropriately if prescribed an NSAID and proton pump inhibitor (PPI), or no longer prescribed NSAID |
| Indicator 2 – GIB02 | Patients 18 years old or over admitted to hospital with a gastro-intestinal bleed prescribed a NSAID and concurrently prescribed an oral anticoagulant (warfarin or a direct oral anticoagulant [DOAC]) | Patients 18 years old or over with a primary care code for GI bleed in 5 years prior to first SMR or start of study, prescribed a NSAID and concurrently prescribed an oral anticoagulant (warfarin or DOAC) in the 3 months prior to SMR | In the 3 months following an SMR, prescriptions were classed as changed appropriately if NSAID and/or oral anticoagulant were no longer co-prescribed |
| Indicator 3 – GIB03 | Patients 18 years old or over admitted to hospital with a gastro-intestinal bleed prescribed an oral anticoagulant (warfarin or DOAC) with an anti-platelet and NOT concurrently prescribed a gastro-protective medicine | Patients 18 years old or over with a primary care code for GI bleed in 5 years prior to first SMR or start of study, prescribed an oral anticoagulant (warfarin or DOAC) with an antiplatelet and not concurrently prescribed a gastro-protective medicine in the 3 months prior to SMR | In the 3 months following an SMR, prescriptions were classed as changed appropriately if PPI was prescribed with an oral anticoagulant and antiplatelet. Alternatively, either the oral anticoagulant or antiplatelet were no longer prescribed |
| Indicator 4 – GIB04 | Patients 18 years old or over admitted to hospital with a gastro-intestinal bleed prescribed aspirin and another anti-platelet and NOT concurrently prescribed a gastro-protective medicine. | Patients 18 years old or over with a primary care code for GI bleed in 5 years prior to first SMR or start of study, prescribed aspirin with another antiplatelet and not concurrently prescribed a gastro-protective medicine in the 3 months prior to SMR | In the 3 months following an SMR, prescriptions were classed as changed appropriately if PPI was prescribed with aspirin and antiplatelet or if aspirin and/or antiplatelet were no longer prescribed |
| Indicator 5 – AKI01 | Patients 18 years old or over admitted to hospital with acute kidney injury concurrently prescribed a non-steroidal anti-inflammatory drug (NSAID), a renin-angiotensin system (RAS) drug, and a diuretic | Not implemented due to data being unavailable. | Not implemented due to data being unavailable. |

| | | | |
|---|---|---|---|
| Indicator 6 – GIBCI | Composite gastrointestinal bleed indicators comprising of unique patients from indicators 1 to 4 | Combination of unique patients from indicators 1-4 | Combination of unique patients from indicators 1-4 |
| Indicator 7 – PAIN01 | Patients 18 years old or over admitted to hospital with respiratory depression, overdose or confusion concurrently prescribed an oral or transdermal opioid and a benzodiazepine, Z-drug, pregabalin or gabapentin | Patients 18 years old or over with a primary care code for respiratory depression in the last five years prior to first SMR or start of study, prescribed an opioid concurrently with a benzodiazepine/ Z-drug/pregabalin/ gabapentin in the 3 months prior to SMR | In the 3 months following an SMR, prescriptions were counted as changed appropriately if opioid no longer concurrently prescribed with a benzodiazepine/Z-drug/ pregabalin/gabapentin |
| Indicator 8 – PAIN02 | Patients 18 years old or over admitted to hospital with constipation prescribed an oral or transdermal opioid and not prescribed a laxative | Patients 18 years old or over with a primary care code for constipation in the last five years prior to first SMR or start of study, prescribed an opioid not concurrently prescribed with a laxative in the 3 months prior to SMR | In the 3 months following an SMR, prescriptions were counted as changed appropriately if a laxative was prescribed with the opioid or if the opioid were no longer prescribed |
| Indicator 9 – PAIN03 | Patients 18 years old or over admitted to hospital with respiratory depression, overdose (accidental poisoning) or confusion currently prescribed an oral or transdermal opioid for more than 3 months | Patients 18 years old or over with a primary care code for respiratory depression in the last five years prior to first SMR or start of study, prescribed an opioid in the 3 months prior to SMR | In the 3 months following an SMR, prescriptions were counted as changed appropriately if the opioid was no longer prescribed |
| Indicator 10 – FRAC01a | Patients 65 years old or over admitted to hospital as a result of a fall prescribed a Z-drug for more than one month | Patients 65 years old or over with a primary care code for a fall in the last five years prior to first SMR or start of study, prescribed a Z-drug in the 3 months prior to SMR | In the 3 months following an SMR, prescriptions were counted as changed appropriately if the Z-drug was no longer prescribed |
| Indicator 11 – FRAC01b | Patients 65 years old or over admitted to hospital with a fracture (hip, colles or humerus) as a result of a fall prescribed a Z-drug for more than one month | Patients 65 years old or over with a primary care code for fracture in the last five years prior to first SMR or start of study, prescribed a Z-drug in the 3 months prior to SMR | In the 3 months following an SMR, prescriptions were counted as changed appropriately if the Z-drug was no longer prescribed |
| Indicator 12 – FRAC02a | Patients 65 years old or over admitted to hospital as a result of a fall prescribed a benzodiazepine for more than one month | Patients 65 years old or over with a primary care code for a fall in the last five years prior to first SMR or start of study, prescribed a benzodiazepine in the 3 months prior to SMR | In the 3 months following an SMR, prescriptions were counted as changed appropriately if the benzodiazepine was no longer prescribed |
| Indicator 13 – FRAC02b | Patients 65 years old or over admitted to hospital with a fracture (hip, colles or | Patients 65 years old or over with a primary care code for fracture in the last five years prior to first SMR or start of study, prescribed | In the 3 months following an SMR, prescriptions were counted as changed |

| | | | |
|---|---|---|---|
| | humerus) as a result of a fall prescribed a benzodiazepine for more than one month | a benzodiazepine in the 3 months prior to SMR | appropriately if the benzodiazepine was no longer prescribed |
| Indicator 14 – FRAC03a | Patients 65 years old or over admitted to hospital as a result of a fall prescribed a benzodiazepine and a Z-drug (not concurrently) for more than one month | Patients 65 years old or over with a primary care code for a fall in the last five years prior to first SMR or start of study, prescribed a benzodiazepine and a Z-drug in the 3 months prior to SMR | In the 3 months following an SMR, prescriptions were counted as changed appropriately if the benzodiazepine or Z-drug was no longer prescribed |
| Indicator 15 – FRAC03b | Patients 65 years old or over admitted to hospital with a fracture (hip, colles or humerus) as a result of a fall prescribed a benzodiazepine and a Z-drug (not concurrently) for more than one month | Patients 65 years old or over with a primary care code for fracture in the last five years prior to first SMR or start of study, prescribed a benzodiazepine and a Z-drug in the 3 months prior to SMR | In the 3 months following an SMR, prescriptions were counted as changed appropriately if the benzodiazepine or Z-drug was no longer prescribed |
| Indicator 16 – RESP01 | Patients 18 years old or over admitted to hospital as an emergency for an exacerbation of asthma prescribed an inhaled Long Acting Beta-agonist (LABA) without an inhaled corticosteroid (ICS) | Patients 18 years old or over with emergency for an exacerbation of asthma in the last five years prior to first SMR or start of study prescribed a LABA but not an ICS in the 3 months prior to SMR | In the 3 months following an SMR, prescriptions were counted as changed appropriately if LABA was prescribed with an ICS or if the LABA was no longer prescribed |
| Indicators 17-20 ACB01-2 | Indicators related to prescribing of medications with moderate or high anticholinergic activity. | Information about prescription of medications with moderate or high anticholinergic activity was not available for analysis in this study. | Not implemented |

In all cases, there was a code for the condition in the last 5 years prior to the SMR and the drug or drugs were prescribed in the 3 months prior to baseline. From these, patients were included if the drug or drugs were prescribed 3 months prior to the SMR and the patient had not died or de-registered following the SMR. All patients were part of the SMR cohort previously extracted. GI=gastro-intestinal bleed; NSAID=non-steroidal anti-inflammatory drug; PPI=proton pump inhibitor; SMR=Structured medication review; DOAC=direct oral anticoagulant; RAS=renin-angiotensin system; LABA=inhaled Long Acting Beta-agonist; ICS=inhaled corticosteroid

**Figure S1.** Incidence of first structured medication reviews by month of observation

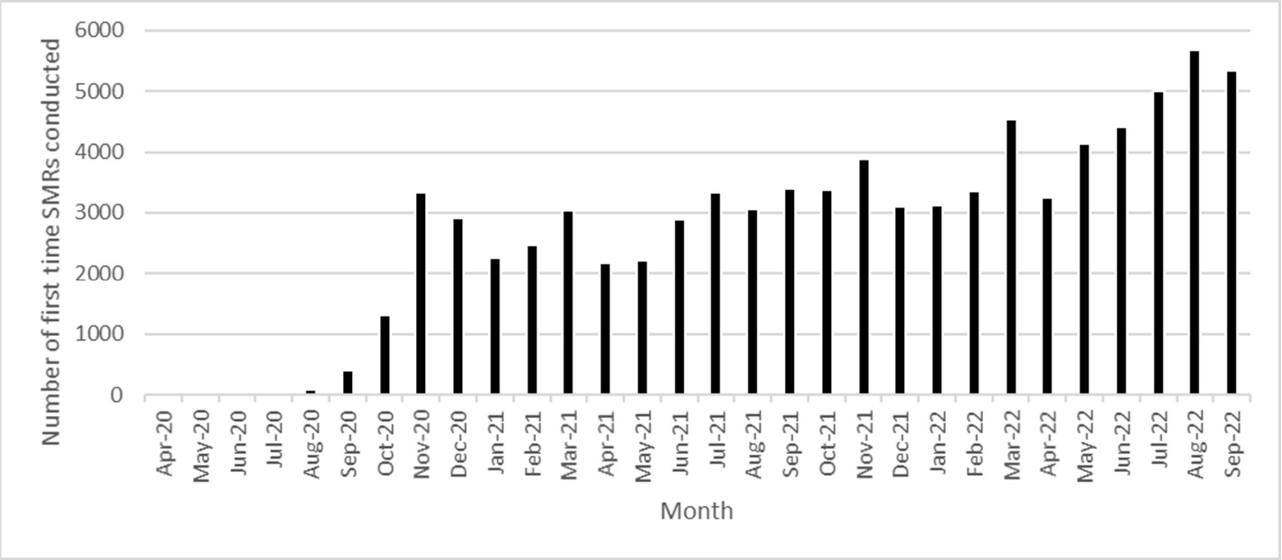

**Table S2.** Multiple long-term conditions recorded at baseline in those patients receiving and not receiving a structured medication review

| **Characteristics** | **Received an SMR** | **Did not receive an SMR** |
|---|---|---|
| Number of long-term conditions (mean (standard deviation) | 5.5 (2.5) | 4.8 (2.4) |
| Addiction | 1,297 (1.6%) | 6,798 (1.2%) |
| Alcohol problem | 3,577 (4.4%) | 22,239 (4.0%) |
| Anxiety / Depression | 35,322 (42.9%) | 215,375 (38.9%) |
| Anorexia | 266 (0.3%) | 1,633 (0.3%) |
| Asthma | 18,684 (22.7%) | 121,734 (22.0%) |
| Atrial fibrillation | 18,213 (22.1%) | 100,628 (18.2%) |
| Blindness | 7,119 (8.7%) | 35,765 (6.5%) |
| Bronchiectasis | 2,769 (3.4%) | 16,225 (2.9%) |
| Cancer | 15,174 (18.4%) | 107,800 (19.5%) |
| Chronic kidney disease | 24,456 (29.7%) | 143,778 (26.0%) |
| Chronic liver disease | 1,318 (1.6%) | 8,473 (1.5%) |
| Chronic pain | 11,432 (13.9%) | 55,540 (10.0%) |
| Connective tissue disorder | 9,916 (12.1%) | 55,005 (9.9%) |
| Constipation | 22,189 (27.0%) | 124,971 (22.6%) |
| Chronic Obstructive Pulmonary Disease | 14,278 (17.4%) | 91,281 (16.5%) |
| Coronary heart disease | 26,097 (31.7%) | 140,555 (25.4%) |
| Dementia | 9,699 (11.8%) | 46,303 (8.4%) |
| Diabetes | 24,773 (30.1%) | 136,675 (24.7%) |
| Diverticulitis | 14,832 (18.0%) | 89,049 (16.1%) |
| Epilepsy | 2,459 (3.0%) | 14,280 (2.6%) |
| Gastric ulcer | 6,404 (7.8%) | 35,085 (6.3%) |
| Hearing loss | 23,960 (29.1%) | 151,105 (27.3%) |
| Heart failure | 11,539 (14.0%) | 61,269 (11.1%) |
| Hypertension | 60,357 (73.4%) | 354,642 (67.7%) |
| Inflammatory bowel disease | 1,770 (2.2%) | 10,377 (1.9%) |
| Irritable bowel syndrome | 8,279 (10.1%) | 52,162 (9.4%) |
| Learning disability | 625 (0.8%) | 1,830 (0.3%) |
| Migraine | 3,563 (4.3%) | 22,113 (4.0%) |
| Multiple sclerosis | 482 (0.6%) | 2,737 (0.5%) |
| Parkinson’s | 1,950 (2.4%) | 10,303 (1.9%) |
| Prostate disorders | 10,995 (33.0%) | 73,899 (31.2%) |
| Psoriasis | 22,886 (27.8%) | 146,530 (26.5%) |
| Peripheral Vascular Disease | 1,686 (2.1%) | 8,228 (1.5%) |
| Schizophrenia/psychosis/bipolar disorder | 1,338 (1.6%) | 7,167 (1.3%) |
| Sinus problems | 2,716 (3.2%) | 17,716 (3.2%) |
| Stroke & transient ischemic attack | 12,116 (14.7%) | 67,828 (12.3%) |
| Thyroids problems | 15,840 (19.3%) | 92,724 (16.8%) |

**Table S3.** Medications of interest recorded at baseline in those patients receiving and not receiving a structured medication review

| Characteristics | Received an SMR | Did not receive an SMR |
|---|---|---|
| **Antihypertensives** | | |
| ACE Inhibitors | 25,079 (30.5%) | 149,864 (27.1%) |
| Alpha blockers | 5,370 (6.5%) | 30,788 (5.6%) |
| Angiotensin II receptor inhibitors | 15,021 (18.3%) | 87,675 (15.8%) |
| Beta Blockers | 28,229 (34.3%) | 158,841 (28.7%) |
| Calcium channel blockers | 26,509 (32.2%) | 164,512 (29.7%) |
| Thiazide and thiazide-like diuretics | 9,092 (11.0%) | 58,861 (10.6%) |
| **Cardiovascular medication** | | |
| Statins | 48,596 (59.1%) | 291,416 (52.7%) |
| Aspirin | 18,294 (22.2%) | 101,067 (18.3%) |
| Other antiplatelets | 26,776 (32.5%) | 145,349 (26.3%) |
| Direct oral anticoagulants | 13,564 (16.5%) | 71,463 (12.9%) |
| Vitamin K agonists | 4,849 (5.9%) | 26,835 (4.8%) |
| **Inhaled medication** | | |
| Inhaled beta agonists | 15,803 (19.2%) | 109,830 (19.8%) |
| Inhaled corticosteroids | 15,377 (18.7%) | 102,567 (18.5%) |
| **Pain medication** | | |
| Non-steroidal anti-inflammatory drugs | 13,654 (16.6%) | 76,324 (13.8%) |
| Opioids | 29,391 (35.7%) | 179,066 (32.4%) |
| **Other medication** | | |
| Antidepressants | 27,636 (33.6%) | 152,684 (27.6%) |
| Benzodiazepines | 5,377 (6.5%) | 32,076 (5.8%) |
| Donepezil | 2,650 (3.2%) | 12,860 (2.3%) |
| Gabapentin | 5,873 (7.1%) | 26,833 (4.8%) |
| Pregabalin | 3,936 (4.8%) | 19,913 (3.6%) |
| Z-drugs | 4,290 (5.2%) | 24,441 (4.4%) |
| Laxatives | 15,947 (19.4%) | 85,196 (15.4%) |
| Proton pump inhibitors | 46,612 (56.6%) | 271,009 (49.0%) |

**Table S4.** Matched cohort baseline characteristics of those receiving and not receiving a structured medication review

| Characteristics | Received an SMR | Did not receive an SMR |
|---|---|---|
| Total cohort population (%) | 71,939 | 71,939 |
| Age at index date (median yrs (p25, p75)) | 77 (71 to 83) | 77 (71 to 83) |
| Sex (% Female) | 30,465 (59.4%) | 30,465 (59.4%) |
| Asian ethnicity | 2462 (3.4%) | 2727 (3.8%) |
| Black ethnicity | 1116 (1.6%) | 1202 (1.7%) |
| Mixed ethnicity | 249 (0.4%) | 279 (0.4%) |
| Other ethnicity | 294 (0.4%) | 348 (0.5%) |
| White ethnicity | 64,014 (89.0%) | 63,180 (87.8%) |
| Unknown ethnicity | 3804 (5.3%) | 4203 (5.8%) |
| BMI (Mean (SD)) | 28.6 (6.2) | 28.3 (5.8) |
| Smoking status | | |
| Active Smoker | 7503 (10.4%) | 7182 (10.0%) |
| Ex-smoker | 30,107 (41.9%) | 28,928 (40.2%) |
| Non-smoker | 33,072 (46.0%) | 35,039 (48.7%) |
| Missing | 1257 (1.8%) | 790 (1.1%) |
| IMD quintile 1 (most deprived) | 11,288 (15.7%) | 10,553 (14.7%) |
| IMD quintile 2 | 12,686 (17.6%) | 12,201 (17.0%) |
| IMD quintile 3 | 14,313 (19.9%) | 13,885 (19.3%) |
| IMD quintile 4 | 14,516 (20.2%) | 14,907 (20.7%) |
| IMD quintile 5 (least deprived) | 13,693 (19.0%) | 14,554 (20.2%) |
| Missing | 5443 (7.6%) | 5839 (8.1%) |
| Living in a rural area | 16,209 (22.5%) | 16,503 (22.9%) |
| Living in an urban area | 52,241 (72.6%) | 51,544 (71.7%) |
| Missing | 3489 (4.9%) | 3892 (5.4%) |
| Systolic blood pressure (mmHg, mean (SD)) | 132.8 (16.9) | 131.1 (16.0) |
| Diastolic blood pressure (mmHg, mean (SD)) | 74.4 (10.0) | 75.0 (9.7) |
| Total cholesterol (mM, mean (SD)) | 4.49 (1.19) | 4.63 (1.18) |
| HDL cholesterol (mM, mean (SD)) | 1.45 (0.45) | 1.50 (0.84) |
| Residing in a care home | 6862 (9.5%) | 947 (1.3%) |
| Frailty | | |
| Fit | 5275 (7.3%) | 11,183 (15.6%) |
| Mild | 21,786 (30.3%) | 28,628 (39.8%) |
| Moderate | 24,535 (34.1%) | 21,056 (29.3%) |
| Severe | 20,343 (28.3%) | 11,072 (15.4%) |
| eFI Score | | |
| <= 0.36 (Not severe) | 56,721 (78.9%) | 64,293 (89.4%) |
| > 0.36 (Severe) | 15,218 (21.1%) | 7647 (10.6%) |
| Polypharmacy | | |
| 0-4 | 2849 (4.0%) | 7725 (10.7%) |
| 5-9 | 18,482 (25.7%) | 26,588 (37.0%) |
| >= 10 | 50,608 (70.4%) | 37,626 (52.3%) |
| Number of MLTC | | |
| 0-1 | 2425 (3.4%) | 4866 (6.8%) |
| 2-3 | 13,466 (18.7%) | 20,312 (28.2%) |
| 4 or more | 56,048 (77.9%) | 46,761 (65.0%) |

SMR=structured medication review; SD=standard deviation; BMI=body mass index; IMD=index of multiple deprivation; MLTC=multiple long-term conditions; eFI=electronic frailty index

**Table S5.** Patient contacts three months before and after the index data in those patients receiving and not receiving a structured medication review

| | 3 months before index date | 3 months after index date | Mean change (95% CI) | Adjusted mean time*intervention difference (95% CI) |
|---|---|---|---|---|
| **(a) All patients contacts** (Median (IQR) contacts per patient (n=patients)) | | | | |
| **Received an SMR** | 4 (2-7); n=71,939 | 5 (3-8); n=71,939 | 1.0 (0.9 to 1.0) | 0.14 (0.13 to 0.16) |
| **Did not receive an SMR** | 3 (1-5); n=71,939 | 3 (1-5); n=71,939 | 0.1 (0.1 to 0.2) | |
| **(b) Patient contacts, excluding SMRs** (Median (IQR) contacts per patient (n=patients)) | | | | |
| **Received an SMR** | 4 (2-7); n=71,939 | 5 (3-8); n=71,939 | 1.0 (0.9 to 1.0) | 0.14 (0.13 to 0.15) |
| **Did not receive an SMR** | 3 (1-5); n=71,939 | 3 (1-5); n=71,939 | 0.1 (0.1 to 0.2) | |
| **(c) Days of patient contacts** (Median (IQR) days of contacts per patient (n=patients)) | | | | |
| **Received an SMR** | 4 (2-6); n=71,939 | 5 (3-7); n=71,939 | 0.9 (0.9 to 1.0) | 0.16 (0.14 to 0.17) |
| **Did not receive an SMR** | 2 (1-5); n=71,939 | 3 (1-5); n=71,939 | 0.1 (0.1 to 0.1) | |
| **(d) Days of patient contacts, excluding SMRs** (Median (IQR) days of contacts per patient (n=patients)) | | | | |
| **Received an SMR** | 4 (2-6); n=71,939 | 4 (3-7); n=71,939 | 0.9 (0.9 to 1.0) | 0.15 (0.14 to 0.17) |
| **Did not receive an SMR** | 2 (1-5); n=71,939 | 3 (1-5); n=71,939 | 0.1 (0.0 to 0.1) | |

Model adjusted for body mass index, ethnicity, index of multiple deprivation, smoking status, care home residence, baseline cholesterol, number of multiple long-term conditions, and electronic frailty index
